# Phenotypic and prognostic insights through unbiased self-supervised learning on kidney histology

**DOI:** 10.1101/2025.06.09.25328891

**Authors:** Krutika Pandit, Nicolas Coudray, Adalberto Claudio Quiros, Aditya Surapaneni, Dhairya Upadhyay, Rami Sesha Vanguri, Daigoro Hirohama, Samer Mohandes, Pascal Schlosser, Heather Thiessen-Philbrook, Yumeng Wen, Chirag R Parikh, Eugene P. Rhee, Sushrut S. Waikar, Insa Schmidt, TRIDENT Study Investigators, Avi Z. Rosenberg, Matthew B. Palmer, Katalin Susztak, Morgan E. Grams, Aristotelis Tsirigos

**Author notes:** co-last authors (Katalin Susztak, Morgan E. Grams, Aristotelis Tsirigos). listed on last page (TRIDENT Study Investigators).

## Abstract

Deep learning methods for image segmentation and classification in histopathology generally utilize supervised learning, relying on manually created labels for model development. Here, we applied a self-supervised framework to characterize kidney histology without the use of pathologist annotations, training on whole slide images to identify histomorphological phenotype clusters (HPCs) and create slide-level vector representations. HPCs developed in the training set were visually consistent when transferred to five diverse internal and external validation sets (1,421 WSIs in total). Specific HPCs were reproducibly associated with slide-level pathologist quantifications, such as interstitial fibrosis (AUC = 0.83). Additionally, hierarchical clustering of tissue patterns revealed patient groups related to kidney function and genotype, and specific HPCs predicted longitudinal kidney function decline. Overall, we demonstrated the translational application of a self-supervised framework to summarize distinct kidney tissue patterns with phenotypic and prognostic relevance.

## Introduction

Advancements in machine learning (ML) have been applied to multiple fields across healthcare.^1–4^ With the increasing availability of large-scale computational resources as well as big data in the form of omics, medical imaging, wearable devices, and electronic health records (EHRs) supporting multi-hospital systems, the implementation of deep learning (DL) can contribute to the integrative use of medical data. The emerging field of computational histopathology exemplifies such an application.^5,6^ It primarily relies on DL methods and the robustness of big data as high-resolution whole slide images (WSIs) to improve the efficiency of tissue characterization, with the ultimate goal of aiding diagnosis and patient outcomes.

DL methods as applied to histopathology can be supervised, wherein models are trained using labels or annotations manually generated by expert pathologists. This approach has shown promising results in the identification of several types of cancers and their subtypes.^7,8^ However, the annotation process is extremely labor-intensive, and pathologist reads can be subjective with high inter-observer variability.^9,10^ Supervised models built on a single pathologist read will perpetuate biases, and thus annotations are often made in parallel by several experts. For example, one study aiming to differentiate between low-grade and high-grade urothelial carcinomas involved the collection of annotations from 21 pathologists over approximately two years of effort.^11^ A study for lung carcinoma classification required a group of 39 surgical pathologists to manually outline boundaries for carcinomatous tissue and non-neoplastic lesions in 4,704 WSIs.^12^ Although the supervised approaches developed in these studies are effective in discerning histological patterns, the meticulous annotation on input images is not easily scalable to complex histopathological processes or specialties with a chronic shortage of experts.^13–15^

Kidney tissue, for example, is especially heterogeneous and many institutions have no dedicated renal pathologists on-site. Subsequently, the few existing studies to characterize kidney pathology using neural networks rely heavily on small patient datasets or animal models.^16–18^

Given the limitations, supervised learning may not be optimal for unbiased and generalizable characterization of histologic patterns in the kidney. As an alternative, self-supervised methods do not require expert labeling and have previously been useful in quantifying multidimensional imaging data. One study, for instance, trained a model on cine cardiac magnetic resonance images using a self-supervised framework and classified patients with preserved or reduced left ventricular ejection fraction.^19^

In this study, we apply a multi-magnification self-supervised learning (SSL) pipeline, initially developed in hematoxylin and eosin (H&E)-stained lung cancer histology, as a method of unbiased quantitative characterization for PAS-stained WSIs of heterogeneous kidney tissue (**Fig. 1**).^20^ Similar to work in speech and text, an effective step in quantification involves the creation of embeddings from smaller components of the input, in this case, image tiles.^21,22^ The embeddings serve as a definition of the component they represent, summarizing its most distinctive features that can later be linked to the broader context of the image, sentence, or audio recording. Each tile embedding can be assigned a histomorphological phenotype cluster (HPC) label via Leiden clustering. This allows for the quantification of every WSI as a vector representation with each vector element accounting for the percentage of image tiles within a given HPC. We hypothesize that distinct, reproducible kidney tissue patterns can be created using a self-supervised deep learning framework without expert labels and can recapitulate pathologist-derived assessments of the entire WSI. We further postulate that the vector representations of the WSIs can be associated with meaningful clinical characteristics, kidney-related outcomes, and genetic parameters.

**Fig 1.**
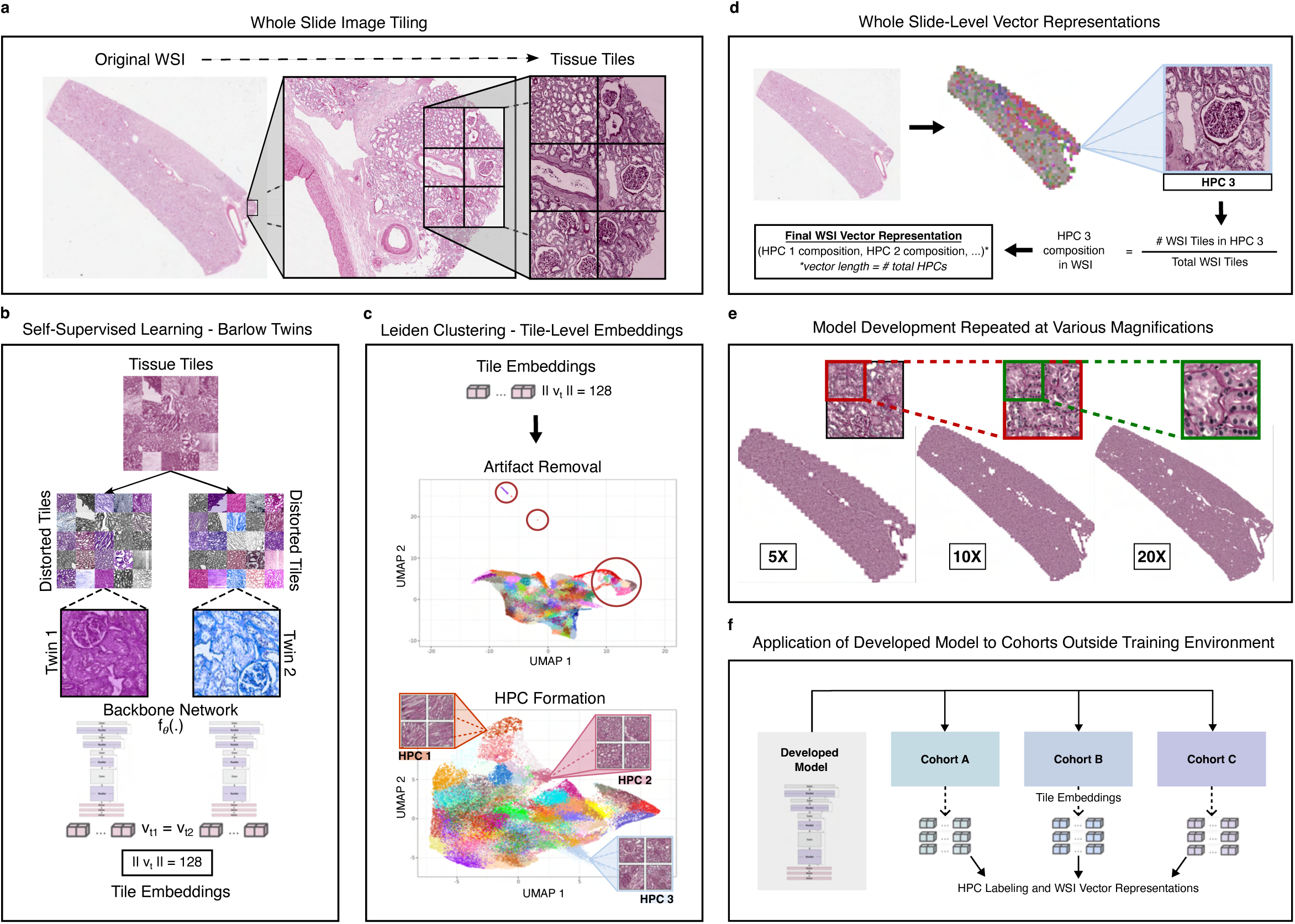
Multi-magnification self-supervised deep learning pipeline. **a,** Whole slide images (WSIs) were processed for tile extraction and stain color normalization. **b,** Self-supervised model backbone f_*θ*_ was trained to create tile embeddings. **c,** Histomorphological Phenotype Clusters (HPCs) were defined using Leiden community detection over a Uniform Manifold Approximation and Projection (UMAP) of tile embeddings. Tile embeddings re-clustered after artifact removal to obtain final HPC labels. **d,** WSI vector representations calculated using percentage of total tissue area in each HPC. **e,** Pipeline (**a-d**) run at 5X, 10X, and 20X magnifications. **f**, Developed model applied to external unseen cohorts to produce tile embeddings, HPC labels, and WSI vector representations.

## Results

### A diverse multi-institutional database of kidney tissue

In total, this study curated a database of 1,421 whole slide images (WSIs) as part of 21 cohorts for robust development and external validation of all findings (**Fig. 2a**). For model development, 739 wedge biopsies were collected during nephrectomies, treated with periodic acid-Schiff (PAS) stains, and digitized at University of Pennsylvania medical institutions (UPenn). The digitized WSIs were randomly grouped into training, validation, and test sets, stratified by estimated glomerular filtration rate (eGFR) at biopsy and history of diabetes. On average, patient age was 61 years old, eGFR was 67 ml/min/1.73 m^2^, 12.5% had 2 or greater dipstick protein, and 35.2% had a history of diabetes (**Fig. 2b**).

**Fig 2.**
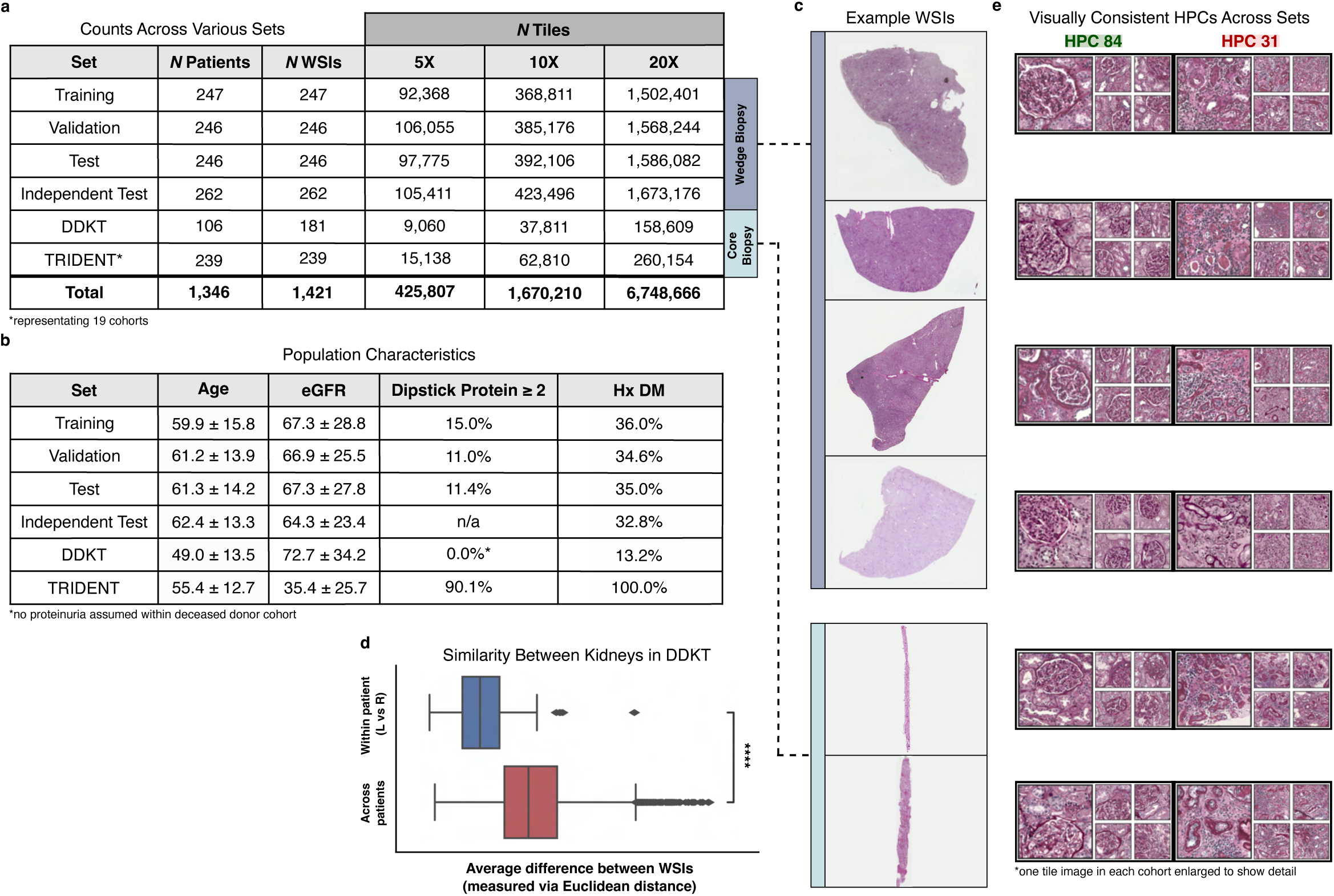
Multi-institutional sources of WSIs used in self-supervised deep learning pipeline. **a,** Number of patients, WSIs, and non-artifactual tiles across training, validation, test, independent test, DDKT, and TRIDENT sets. **b**, Age, eGFR, dipstick protein (≥ 2), and history of diabetes across sets. **c,** Example WSIs from each set colored by type of biopsy (wedge or core). **e,** Representative tiles from HPCs 84 and 31. **d,** Average within- and across-patient Euclidean distances between WSI vector representations plotted in set with left and right kidney WSIs (DDKT, *p*=4.5e-23).

Three additional cohorts of PAS-stained WSIs were acquired one year after the initial development set – an independent test set, a deceased donor kidney transplantation (DDKT) set, and a set of clinical biopsies from the Transformative Research In DiabEtic NephropaThy (TRIDENT) study (representing a collection of 19 cohorts, **Fig. 2c**). The independent test set included 262 WSIs derived from wedge biopsies collected from nephrectomies at UPenn and digitized at New York University (NYU). The DDKT set was derived at Johns Hopkins University (JHU) from pre-implantation core biopsies consisting of 106 patients, 75 of whom had left and right kidney samples (**Fig. 2d**). The TRIDENT set consisted of 239 WSIs of core biopsies from patients with diabetic nephropathy. All WSIs were divided into image tiles at 5X (452 x 452- µm/tile), 10X (226 x 226-µm/tile), and 20X (113 x 113-µm/tile) magnifications resulting in 425,807, 1,670,210, and 6,748,666 image tiles, respectively.

### An unbiased atlas of kidney histology developed through self-supervised learning

Next, tile embeddings were generated using a self-supervised deep learning framework. To avoid overfitting, epochs were visualized against loss in the training and validation sets during model development. When no further improvement was seen in validation loss, training was stopped at epoch 30 (**Supplementary Fig. 1**). At this stage, each tile had an embedding of length 128. Using tile embeddings from the training set, two rounds of Leiden clustering were performed.

The first round generated a large number of HPCs for maximum differentiation of tissue patterns. Representative tiles were visually examined in the training set to identify artifact HPCs (e.g., blurry, edge, and non-tissue tiles; **Supplementary Fig. 2a**). Labels from round one of Leiden clustering were also assigned to external sets to remove all artifact image tiles. A second round of clustering was performed on the remaining tile embeddings to form non-artifact HPCs in the training set. The optimal number of HPCs was determined using a connectivity metric called disruption score (**Supplementary Fig. 2b**). Overall, the pipeline produced an atlas of 77, 100, and 99 HPCs at 5X, 10X, and 20X magnifications, respectively (**Supplementary Fig. 2c**).

Non-artifact HPC labels were transferred to validation, test, independent test, DDKT, and TRIDENT sets. Visual inspection of HPCs by a pathologist revealed that histological patterns were consistent across all sets (**Fig. 2e**, 10X example). Additionally, we created a tile-set matching exercise to quantitatively assess visual consistency of HPCs across the training, validation, test, independent test, DDKT, and TRIDENT sets. Each question in the exercise displayed tile samples of a particular HPC (e.g. HPC 1) from five out of the six sets. The user was asked to choose a matching sample of tiles from the sixth set out of three options. We surveyed 21 people (nine nephrology experts/kidney pathologists; 12 non-experts). Average score was 94.2.0% (SD = 11.7%) among experts and 90.0% (SD = 13.1%) among non-experts. Representative tiles from all HPCs and sets at 5X, 10X, and 20X can be viewed here – https://genome.med.nyu.edu/public/pmedlab/SSL_kidney/representative_tiles.

Compositional vector representations were calculated for each WSI. Each vector element corresponded to the percentage of total tissue area assigned to a given HPC, also referred to as HPC composition. Average presence of HPCs was visualized as a histogram by set (**Supplementary Fig. 3**). In the DDKT dataset, it was confirmed that the Euclidean distance between WSI vector representations were more similar within-patient (left versus right kidney) compared to across-patient (*p* = 4.5e-23).

### Slide-level associations of HPC composition with pathologist quantification

Focusing on 10X magnification as an example (5X and 20X magnifications included as **Supplementary Fig. 4**), we observed statistically significant correlations adjusted for multiple hypothesis testing using the Bonferroni method between HPC composition and slide-level pathologist quantification in the training set (**Fig. 3a**). These correlations were consistent across validation, test, independent test, and TRIDENT sets (**Fig. 3b**). For example, HPCs correlated with interstitial fibrosis were also correlated in external sets and demonstrated fibrotic tissue patterns on visual inspection by an expert pathologist. Three WSIs from the test set were visualized with tiles classified in fibrosis-associated HPCs highlighted in red (**Fig. 3c**). The corresponding pathologist annotations for interstitial fibrosis displayed agreement between regions represented by fibrosis-associated HPCs and regions highlighted as fibrotic by the expert. A multiple instance learning (MIL) model (CLAM) using HPC composition as features to predict binarized percent interstitial fibrosis (>5%) developed in the training set and optimized in the validation set demonstrated an AUC of 0.79 and 0.83 in the test and independent test sets, respectively (**Fig. 3d**).^23^

**Fig 3.**
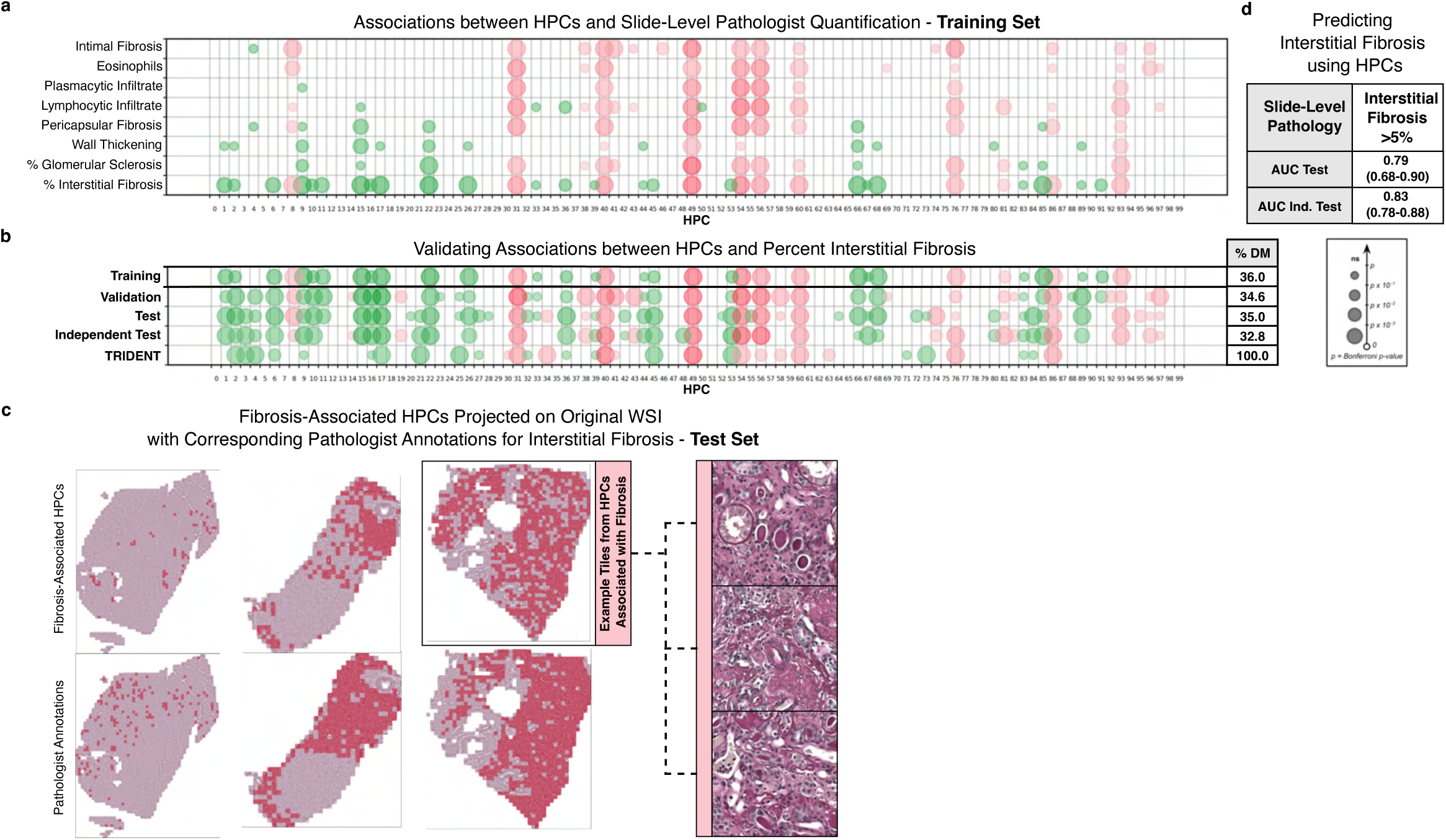
Analysis of HPC composition against pathologist quantification. **a,** HPC composition was correlated against slide-level pathologist quantifications in the training set. Only Bonferroni significant associations shown. Circle size increases with significance. Red circles indicate positive associations with pathology; green circles indicate negative associations with pathology. **b,** Analysis was repeated in the validation, test, independent test, and TRIDENT sets for HPCs significantly correlated with percent interstitial fibrosis in the training set. For HPC representation in the different sets, see Supplementary Fig. 3. **c,** Tiles classified in HPCs significantly associated with percent interstitial fibrosis were highlighted on three WSIs from the test set in red. Given are the corresponding pathologist annotations for interstitial fibrosis. For the WSI with the highest prediction, three example tiles from HPCs significantly associated with percent interstitial fibrosis are shown. **d,** MIL model was trained to predict binarized percent interstitial fibrosis (>5%) using proportion of patients’ tiles in each HPC. AUC and standard error shown for performance in the test and independent test sets.

To assess the current SSL algorithm against off-the-shelf publicly-available models, a comparative analysis was conducted against pre-trained ResNet-50 and UNI^24–26^. Tile embeddings generated from each pre-trained network for the training, validation, and test sets were clustered via Leiden. The resulting Leiden clusters were associated with slide-level percent interstitial fibrosis. HPL-derived Leiden clusters showed significantly stronger associations with interstitial fibrosis as compared to those derived from ResNet-50 and UNI (**Supplementary Fig. 5a**). Furthermore, UNI-derived Leiden clusters projected on to TRIDENT PAS- and H&E-stained tissue tiles (collected from WSIs created from the same tissue core) revealed stain-specific and patient-specific batch effects (**Supplementary Fig. 5b**).

### HPC composition is prognostic of kidney function decline

In the TRIDENT set, which was followed longitudinally for the development of end-stage kidney disease (ESKD) or chronic kidney disease (CKD) progression (a greater than 40 percent drop in eGFR during a maximum follow-up period of 1,200 days post biopsy), select HPCs associated with interstitial fibrosis were also associated with kidney function decline. For instance, patients in the highest tertile of composition for HPC 49 (annotated “global sclerosis (solidification) in the context of interstitial fibrosis and tubular atrophy (IFTA)” by a blinded pathologist) had greater kidney function decline, whereas patients in the highest tertile of composition for HPC 53 (annotated “histologically unremarkable proximal tubules” by a blinded pathologist) had less kidney function decline (**Fig. 4a**, **Fig. 4b**). Previous studies have demonstrated a strong link between kidney function decline and the presence of IFTA.^27,28^

**Fig 4.**
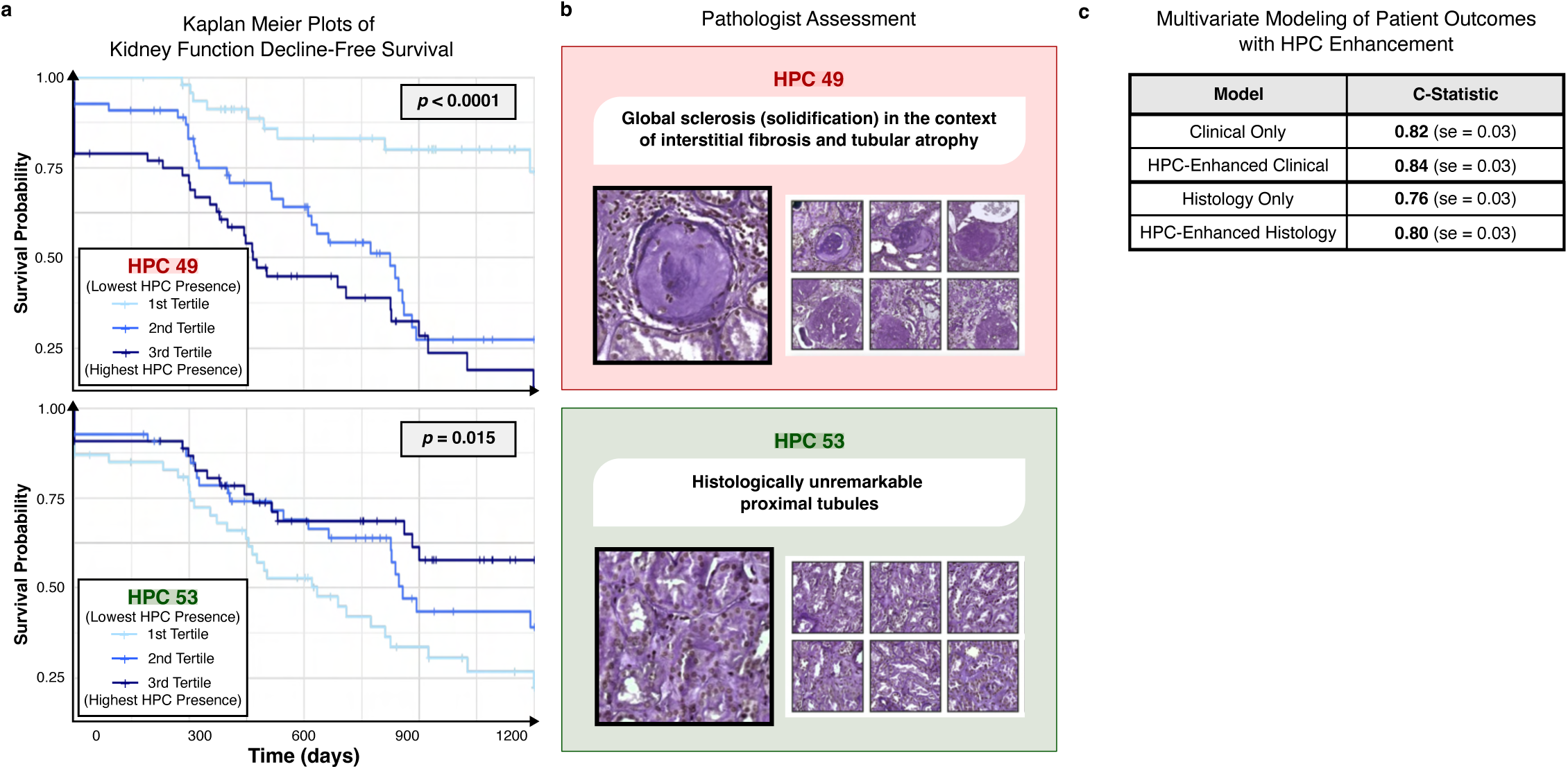
Modeling kidney-specific outcomes using HPC presence in WSI. **a,** Cumulative survival probabilities for kidney-specific patient outcomes plotted for tertiles of prognostic HPCs in the TRIDENT set. Primary outcome was defined as the first instance of end-stage kidney disease (ESKD; defined as dialysis over 3+ months or kidney transplant) or CKD progression (defined as a drop of at least 40 percent in eGFR from baseline) during a mean follow-up period of 468 days post biopsy. HPC 49 highlighted in red; HPC 31 highlighted in green. **b,** Representative tiles from HPCs 49 and 53 were labeled by an expert pathologist in a blind experiment. **c,** Multivariate Cox proportional hazards models were trained using only clinical variables (age, sex, ACR, eGFR) and only histology variables (percent interstitial fibrosis, RPS score, epithelial hyperplasia) to predict primary outcome. Analyses were repeated in combination with four HPCs (34, 40, 49, 86) selected based on non-zero coefficients in a LASSO regression analysis.

The ability of HPC composition to predict kidney function decline was further investigated using multivariate Cox proportional hazards models in the TRIDENT set (**Fig. 4c**). In 182 patients with relevant longitudinal and clinical data, a model built on established kidney-related clinical risk factors – age, sex, albumin-to-creatinine ratio (ACR), and eGFR – was used to predict ESKD or CKD progression (c-statistic = 0.82).^29,30^ Discrimination of the model for kidney function decline improved when four HPCs (34, 40, 49, 86) selected based on univariate analyses (*p* < 0.1) and non-zero coefficients in a Lease Absolute Shrinkage and Selection Operator (LASSO) regression were added to the clinical model (c-statistic = 0.84). Similarly, a second baseline model was established using slide-level histology known to be associated with eGFR decline – interstitial fibrosis, Renal Pathology Society (RPS) score, and epithelial hyperplasia (*N* = 177, c-statistic = 0.76).^31,32^ Discrimination improved when analysis was repeated in conjunction with the four selected HPCs (c-statistic = 0.80).

### Hierarchical clustering of HPC composition classifies patients into distinct groups marked by clinical parameters and kidney genotype

We then hypothesized that HPC composition can be used to group patients by histologic similarity. To this end, hierarchical clustering was applied at the patient level using Euclidean distance between the compositional WSI vector representations to reveal five remarkably distinct patient groups (**Fig. 5a**). Next, for each patient group, a marker HPC was defined as the HPC with the greatest difference in composition between groups. Visualization of representative tiles from marker HPCs revealed varying tubular and medullar morphologies (**Fig. 5b**). The marker HPC for patient group 4, HPC 8, was further assessed against clinical variables due to its distinctly medullar composition. There was minimal association between HPC 8 and eGFR and proteinuria, although relatively few patients had quantitative assessment of proteinuria (**Supplementary Fig. 6**).

**Fig 5.**
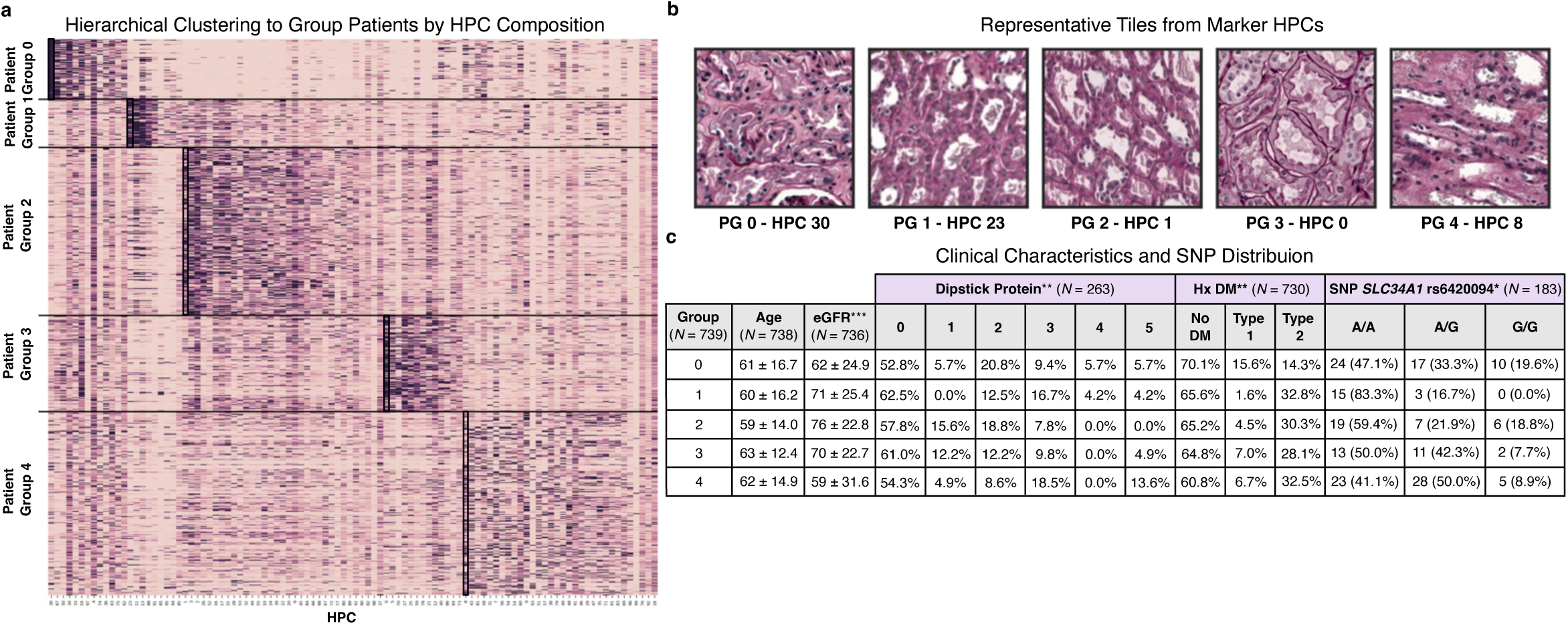
Formation of distinct patient groups based on HPC composition. **a,** Hierarchical clustering of patients performed using Euclidean distances between WSI vector representations. Marker HPCs for five patient groups (PG) were defined using differential HPC composition highlighted in black. **b,** One representative tile shown from each marker HPC. **c,** Differences in clinical characteristics and genetic variation analyzed using ANOVA and Chi-squared tests.

Differences in patient characteristics, including three directly genotyped single-nucleotide polymorphisms (SNPs) of interest available in a subset of the cohort, were then assessed between patient groups (*N* = 183, **Fig. 5c**). Patient Group 4 displayed significantly lower eGFR (*p* = 2.0e-10) and higher dipstick protein levels (*p* = 0.008) than the other four groups. Patient Group 0 contained a relatively higher percentage of patients with Type 1 diabetes (*p* = 0.006). Patient Group 1 demonstrated differences in allele frequency of rs6420094 in the *SLC34A1* gene, a gene that has previously been categorized in a targeted CRISPR/Cas9 study as indispensable to maintain kidney function.^33^ It has also been implicated in GWAS meta-analyses for nephrolithiasis, osteoporosis, and the mediation of inorganic phosphate reuptake as an encoder for the type IIa Na/P_i_ co-transporter in the tubular region which may contain risk variants for renal tumors and chronic kidney disease.^34^

Additionally, a comprehensive analysis of patient groups defined using HPC composition revealed significant differences in history of HTN, kidney health measures, and slide-level histology annotations (**Supplementary Table. 1**). This analysis was repeated within patient groups defined using HPC composition from all available magnifications (5X, 10X, and 20X) resulting in significant differences in BMI, kidney health measures, and slide-level histology annotations (**Supplementary Table. 2**).

## Methods

### Data acquisition and preprocessing

Non-cancerous kidney tissue from 739 participants was collected during nephrectomies and digitized at UPenn using the Hamamatsu NanoZoomer S360 at 40X magnification. For model development, participants were split into three equal sets stratified by history of diabetes and eGFR – training set (*N* = 247), validation set (*N* = 246), and test set (*N* = 246). Three additional cohorts of PAS-stained WSIs were received one year after the development set - 262 samples from UPenn (digitized at NYU using the Leica Biosystems Aperio AT2 at 40X magnification) called the independent test set, 181 samples including 75 left and right pairs of kidney samples called the DDKT set (digitized at JHU using the Leica Biosystems Aperio AT2 at 40X magnification), and 239 samples from a diabetic nephropathy study combining efforts from 19 institutions called the TRIDENT set (digitized at UPenn using the Hamamatsu NanoZoomer S360 at 40X magnification).^35^ PAS-staining in all WSIs was confirmed upon careful evaluation by the pathologist authors.

All WSIs were divided into non-overlapping tiles with dimensions of 224 x 224 pixels using the publicly available DeepPATH pipeline for preprocessing (**Fig. 1a**).^8^ Tiles with over 50 percent background were excluded. As a means to address variation in staining depth and quality, Reinhard’s method was used for stain normalization aiming to match the mean and standard deviation of 100 randomly selected tiles from the UPenn dataset.^36^ Tiles were generated from all WSIs at 5X, 10X, and 20X magnifications with each pixel representing 2.016µm, 1.008µm, and 0.504µm of tissue respectively.

### Model development and artifact removal

A self-supervised learning framework was used to capture and convert morphological characteristics in WSIs into vector representations.^20^ First, the BarlowTwins algorithm which aims to create embeddings for the input image was applied to each tile in the training set (**Fig. 1b**).^37^

The neural network architecture is composed of several ResNet layers and a self-attention layer. Two altered (e.g. rotated, recolored) versions of the input tile were passed through identical neural networks, each producing an output vector of length 128. To make the embeddings invariant to distortions and reduce redundancy between components of the embeddings, the objective function in the BarlowTwins algorithm measures the cross-correlation matrix of the two output vectors making it as close to an identity matrix as possible. Learning was monitored using loss in training and validation sets calculated at the end of each epoch, a hyperparameter that specifies the number of times the neural network is exposed to the full training set. Training was stopped at epoch 30 when no further decrease in validation loss was detected. Vector representations for tiles in the validation, test, independent test, DDKT, and TRIDENT sets were generated using the trained SSL backbone at epoch 30.

Next, Leiden community detection defined a nearest neighbor graph of the vector representations to find HPCs of visually similar tiles (**Fig. 1c**).^38^ This step was conducted as two rounds at varying resolutions, a hyperparameter used to specify the desired number of HPCs resulting from clustering. Round one of clustering used a high resolution for visual identification (in training set) and removal (in all sets) of artifact (HPCs containing blurriness, edges, or non-tissue material). Round two aimed to create HPCs from the remaining kidney tissue tiles at an optimal resolution. The optimal resolution or number of HPCs was determined using a connectivity metric called disruption score that quantifies tightness and separation of clusters. This score was calculated at each resolution as the sum of distances between each tile within an HPC and its corresponding centroid averaged across all HPCs.

HPCs were defined at various resolutions in the training set, and the resulting labels were applied to tiles in the validation set. Disruption score was plotted for all resolutions in the validation set, and the number of HPCs at the elbow was chosen as optimal. All above steps were conducted at 5X, 10X, and 20X magnifications; the optimal resolutions were determined to be 5.0, 5.0, and 3.5, respectively.

### WSI quantification and associations with expert-annotated pathology

Once Leiden labels were established, representative tiles from each HPC were visually inspected for consistently across training, validation, test, independent test, DDKT, and TRIDENT sets by an expert pathologist. Visual consistency of HPCs across sets was objectified using a tile- set matching exercise consisting of ten multiple choice questions. Each question provided tile samples of a particular HPC (e.g. HPC 1) from five out of the six sets. Each tile sample contained five tissue tiles. Users were asked to choose a matching sample of tiles from the sixth set, given three answer choices. The two remaining incorrect answer choices were tile samples from other HPCs (e.g. non-HPC 1). Participants ranged from nephrology experts/kidney pathologists to non-experts.

A compositional vector was calculated to summarize the morphological findings in a WSI, each vector element being the percentage of a given HPC with respect to the total tissue area (**Fig. 1d**). Original WSIs were tiled at all magnifications (**Fig. 1e**). For the model developed at 10X magnification, the Euclidean distances between within-patient and outside-patient WSI vector representations were calculated for comparison in the DDKT set.

HPC composition within the vector representations was correlated with slide-level expert quantification. Spearman correlations were validated across all sets with relevant clinical data using a Bonferroni threshold. Next, WSI vector representations were used as input for an MIL model to predict binned level of interstitial fibrosisA positive instance of interstitial fibrosis was defined as being above median percent interstitial fibrosis (5%) in the training, validation, and test sets. .^39^ Performance of developed model was assessed on hidden test and independent test sets.

Performance of the current SSL model was compared against off-the-shelf models - ResNet-50 (pre-trained on ImageNet) and UNI (pre-trained on ∼100k WSIs of diverse H&E-stained tissue). Embeddings for tiles from the training, validation, and test sets were extrapolated from each model and cluster labels were generated using Leiden. Strength of associations between each set of Leiden clusters and percent interstitial fibrosis was used as the metric of comparison and visualized across models as a boxplot. UNI-derived Leiden clusters were further analyzed within the TRIDENT set containing, both, PAS and H&E WSIs derived from the same tissue core.

### Predictive modeling of kidney function decline using HPC composition

In the longitudinal TRIDENT cohort, WSIs were split into tertiles based on HPC composition and plotted as Kaplan Meier curves to distinguish varying rates of kidney function decline, defined as the development of ESKD or CKD progression (40 percent drop in eGFR during a maximum follow-up period of 1,200 days after biopsy). Two baseline Cox proportional hazards models predicting kidney function decline were developed using known clinical risk factors (age, sex, ACR, eGFR) and slide-level histology measures (percent interstitial fibrosis, RPS score, epithelial hyperplasia). HPCs were selected based on univariate analyses (*p* < 0.1) and subsequent LASSO regression. The sparsity parameter λ was set to 0.14 in the LASSO analysis to select four HPCs. Cox analyses were repeated in combination with the four selected HPCs (HPC-enhanced clinical, HPC-enhanced histology).

### Hierarchical clustering of patients

Patients from training, validation, and test sets were hierarchically clustered based on Euclidean distance between WSI compositional vectors to create five groups. Optimal number of patient groups was determined using disruption score. For each of the five patient groups, average HPC presence was calculated within-group and outside-group. The HPC with highest in- group and out-group differences was chosen as the marker HPC. The marker HPC for patient group 4 for further analyzed for associations with eGFR and proteinuria.

Patient groups were also analyzed for differences in clinical characteristics such as eGFR, urine dipstick, medical history, and slide-level histology annotations. Continuous and percent variables were compared using ANOVA and chi-squared analyses (*p* < 0.05), respectively. Additionally, a subset of patients had available genotyping for the following SNPs - *UMOD* rs12917707, *SLC34A1* rs6420094, and *SLC7A9* rs12460876. Variation in alleles between the five patient groups was assessed using a genotypic genetic model.

## Discussion

The current translational application of a self-supervised deep learning framework to characterize kidney histology highlights that *(1)* SSL might obviate the need for time-consuming pathologist annotations on large datasets for model development compared to a supervised approach, *(2)* tissue patterns detected from SSL in a training set are reproducible in external cohorts, both visually and vis-a-vis expert quantification, *(3)* histological patterns detected using SSL can be used to model long-term kidney outcomes such as ESKD and eGFR decline, *(4)* WSI quantification resulting from SSL can be used to create patient groups with distinctive kidney function and possible links to genetic variation, and *(5)* this methodology for characterization can be applied across histological domains with varying tissue complexity.

SSL circumvents the need for pathologist annotations in model development and in this study enabled use of a relatively large dataset (1,421 WSIs). In contrast, supervised tasks require time-consuming annotations, which often severely limit the size of the dataset. In histopathology, the annotation process is further complicated by the relative paucity of pathologists. Many institutions lack even a single on-site kidney pathologist. Previous supervised studies characterizing histopathologic structures in the kidney exemplify this barrier. One study required three board-certified pathologists to build a model to quantify glomerulosclerosis from 83 donor kidneys.^28^ Another study required categorical labels created by five kidney pathologists to estimate IFTA on 67 WSIs.^40^ Our study proposes an effective SSL approach as a scalable alternative that does not require manual labeling.

The use of large, unselected datasets for model development may improve the transferability of learned features and minimize the impact of technical factors such as differences in depth of biopsy, stain penetration, and methods for digitization.^40–42^ In our study, WSIs in the training set were derived from nephrectomies and thus represent pathologies commonly seen in the general population (e.g., diabetic changes, hypertensive changes).^26^ The developed models performed well in external validation sets, which encompassed clinically diverse samples from over 20 institutions. Moreover, the size of biopsies varied between development and validation WSIs: the development sets were from wedge biopsies (relatively large tissue size) and most external validation samples were from needle biopsies (relatively small tissue size). Thus, an SSL approach that enables training on large datasets may help optimize generalizability.

The learned features of an SSL model (e.g., tile embeddings or WSI vector representation) can be used in various downstream tasks. In this study, we used WSI vector representations to identify distinct patient phenotypes, creating groups of patients that differed by level of kidney function or genetic variants. A similar approach has been employed using EHR data or genetic data.^43–45^ However, tile embeddings could also be directly used as input features for supervised prediction in attention-based methods such as multiple instance learning.^46–49^ Thus, the output of an SSL model may be adaptable and tile-level clustering may not be necessary for all downstream tasks.

The vision for AI-augmented healthcare is to produce fair and interpretable insights that improve patient health.^50–52^ Clinical decision-making tools powered by data-driven models have been successfully deployed in local settings. At NYU, a deep neural network to predict the likelihood of COVID-19 based on chest x-rays was tested in clinical practice.^53^ With further validation, our model could provide prognostic information for two prominent outcomes of interest for kidney function – ESKD and eGFR decline – using digitized histology inputs. Model output could also be used to provide visual and quantifiable insights for clinicians through the projection of HPCs onto the original WSI. With dissemination to other institutions, SSL-derived approaches such as ours could increase efficiency for kidney pathologists, standardize scoring systems, and identify novel prognostic tissue patterns (**Fig. 6**).

**Fig 6.**
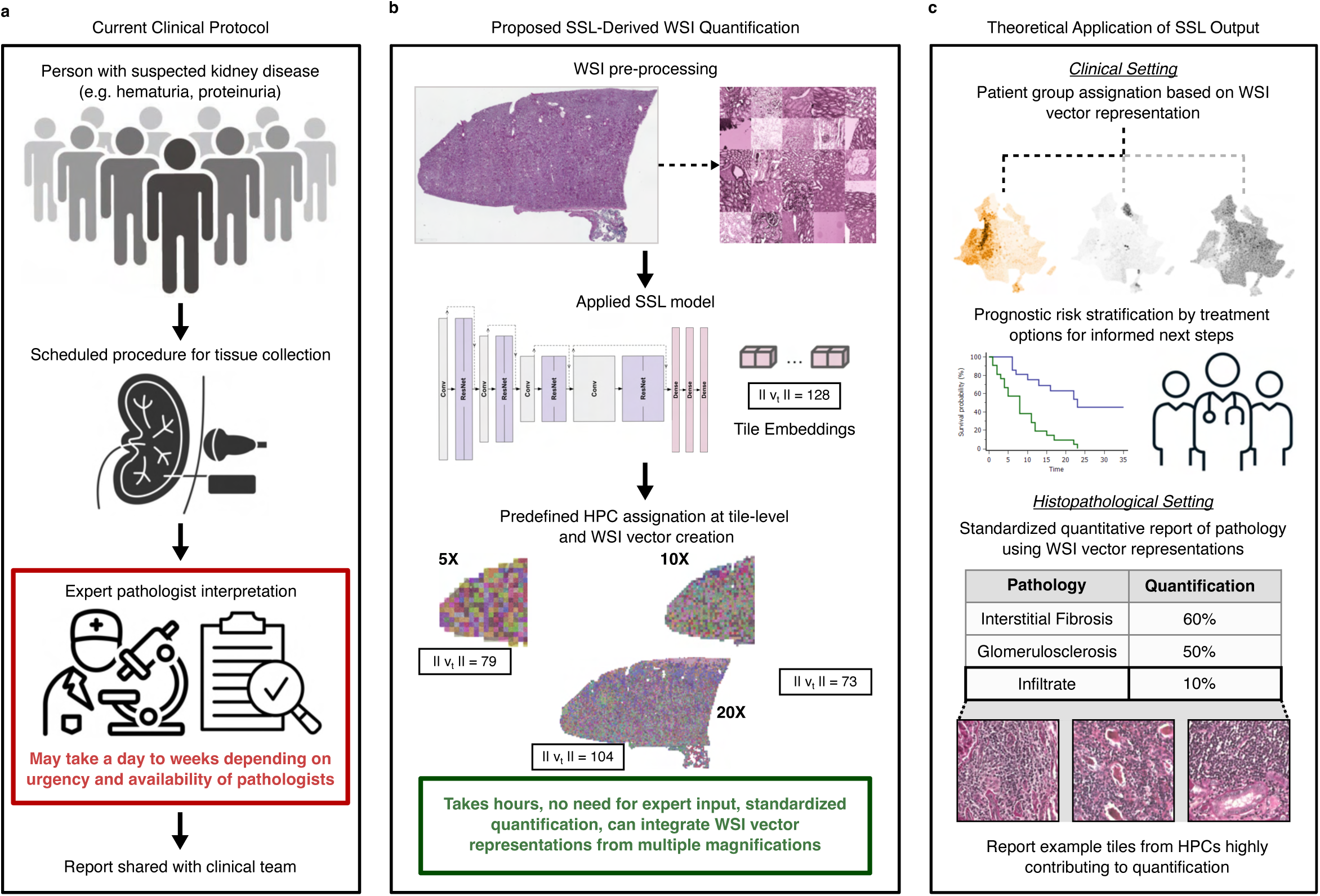
Clinical and histopathological use of WSI quantification by SSL. **a,** Outline of current clinical protocol for identification of patients suspected of having kidney disease, collection of kidney tissue, and evaluation by expert pathologist. **b,** Outline of proposed protocol incorporating outputs from SSL framework for expedited processing and quantification of kidney tissue patterns. **c,** Proposed clinical and histopathologic applications for quantified kidney histology.

This study has limitations. First, the training set does not incorporate rare causes of kidney disease and, thus, does not represent the full disease spectrum encountered in nephrological practice. Future work should prioritize the capture of a wider range of biopsies from patients with rarer etiologies as well as from transplant recipients. This could facilitate the creation of a “databank” of diverse WSI vector representations allowing for downstream supervised assessments (such as MIL) and further unsupervised clustering. Second, pre-training models on non-PAS-stained WSIs would greatly expand the relevance of this pipeline to a wider range of pathologies detected by stains such as H&E. Third, the role of tile magnification should be further explored. Smaller features such as eosinophil infiltration might be better quantified at higher magnifications, whereas contextualized structures such as pericapsular fibrosis might be better detected at lower magnifications. Fourth, the tradeoff between choosing a small number of clusters to support human interpretability and choosing a large number of clusters to capture a wide range of distinct morphologies can be further explored. Lastly, the current pipeline does not incorporate pre-training on publicly available, non-histology specific sources such as ImageNet. While pre-training is computationally intensive, there may be potential advantages of pre-training such as faster convergence for fine-tuning steps as compared to models developed without pre-training.^54^

Overall, we have shown that a self-supervised DL framework can be applied to kidney histopathology to provide detailed quantification of tissue patterns without the need for pathologist labeling. This approach allows for the use of increasingly large datasets which, when comprised of data from clinically and geographically diverse sources, increases the utility and generalizability of DL methods. Histological patterns quantified from these methods can identify phenotypically and genotypically meaningful groups of patients and predict adverse events. With additional development and validation, these tissue patterns might be implemented as standardized scoring for improved pathologist efficiency and clinician interpretation.

## Code Availability

The HPL pipeline can be replicated using the following GitHub repository - https://github.com/AdalbertoCq/Histomorphological-Phenotype-Learning.

Models from the current study trained on kidney tissue tiles of 5X, 10X, and 20X magnification are available here - https://github.com/AdalbertoCq/Histomorphological-Phenotype-Learning/blob/master/README.md#other-pretrained-models.

## Data Availability

Access to training data (whole slide images) used in the present study may be available upon request. The HPL pipeline can be replicated using the following GitHub repository - https://github.com/AdalbertoCq/Histomorphological-Phenotype-Learning. Models from the current study trained on kidney tissue tiles of 5X, 10X, and 20X magnification are available here - https://github.com/AdalbertoCq/Histomorphological-Phenotype-Learning/blob/master/README.md#other-pretrained-models.

## TRIDENT Study Investigators

| Site | Name |
| --- | --- |
| UAMS | Manisha Singh |
| Einstein | Michael Ross |
| GA | James Tumlin |
| MSSM | Kirk Campbell |
| UNC | Amy Mottl |
| UNM | Christos Argyropoulos |
| Northwestern | Tamara Isakova |
| OSU | Salem Almaani |
| OHSU | Rupali Avasare |
| Stanford | Richard Lafayette |
| UVA | Julia Scialla |
| Yale | Randy Luciano |
| UTHSA | Shweta Bansal |
| Univ of Arizona | Frank Brosius |
| Baylor | Ankit Mehta |
| Miami | Oliver Lenz |
| LVHN | Nelson Kopyt |
| Columbia | Piettro Canetta |
| Michigan | Matthias Kretzler |
| Metro | Jeffrey Schelling |

## Notes

### Competing Interest Statement

Aristotelis Tsirigos is a cofounder of Imagenomix.

### Clinical Protocols

https://github.com/AdalbertoCq/Histomorphological-Phenotype-Learning

### Funding Statement

This was an NIH-funded study.

### Author Declarations

Ethics committee/IRB of the University of Pennsylvania gave ethical approval for this work. Ethics committee/IRB of Johns Hopkins University gave ethical approval for this work.

