## Supplemental Figures for "Phenotypic and prognostic insights through unbiased self-supervised learning on kidney histology"

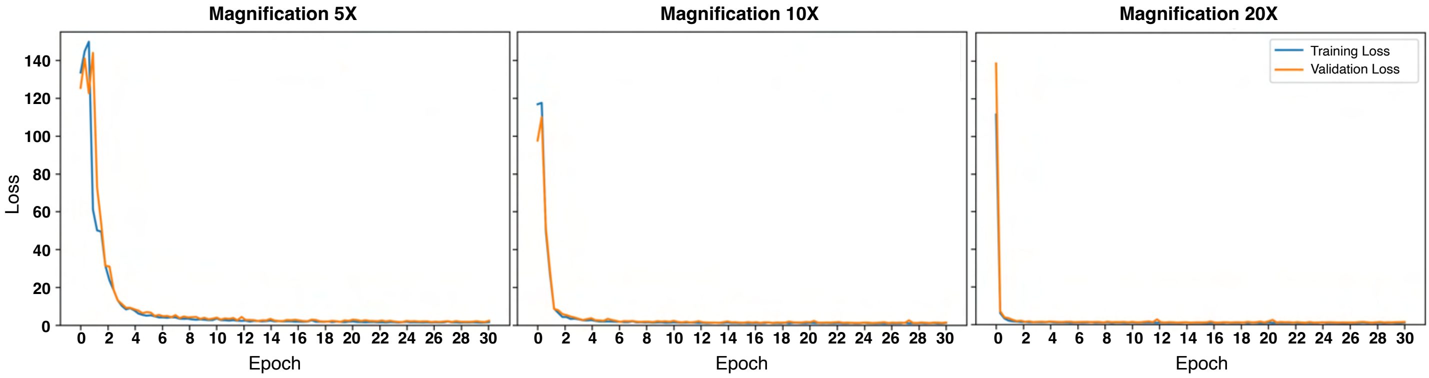


**Supplementary Fig. 1 | Training and validation loss during BarlowTwins model development.** Loss was monitored during SSL training to prevent overfitting to training data. Backbone network at epoch 30 was used to create WSI vector representations at the tile level in training, validation, test, independent test, DDKT, and TRIDENT sets.

**a**

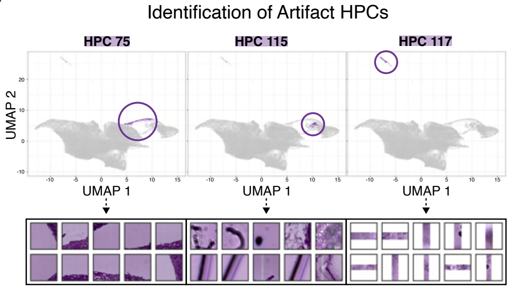


**b**

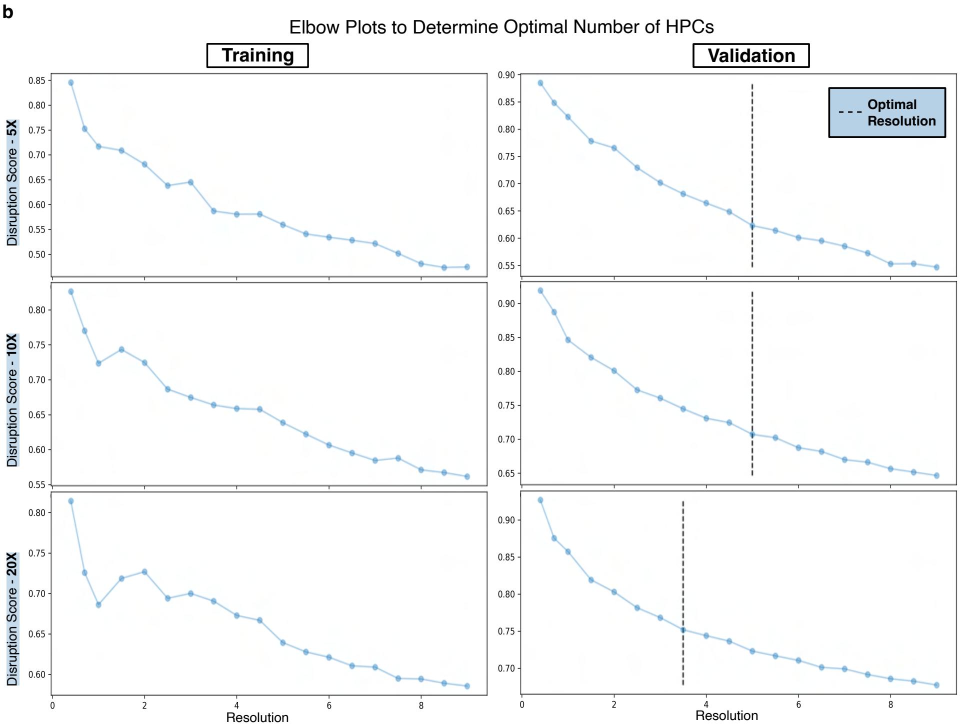


**c**

Number of HPCs by Magnification

| Magnification | Optimal Resolution | Number of HPCs |
|---|---|---|
| 5X | 5.0 | 77 |
| 10X | 5.0 | 100 |
| 20X | 3.5 | 99 |

**Supplementary Fig. 2 | Artifact removal and identification of optimal Leiden resolution. a,** Tile embeddings plotted as UMAPs colored by artifact HPCs identified using round one of Leiden clustering at high resolution (9.0). Representative tiles for artifact HPCs 75, 115, and 117 shown. **b,** Disruption score plotted against increasing Leiden resolution in round two of Leiden clustering. Elbow identified in validation set as optimal resolution to form non-artifactual kidney tissue HPCs. **c,** Number of clusters resulting from optimal resolution at 5X, 10X, and 20X magnifications.

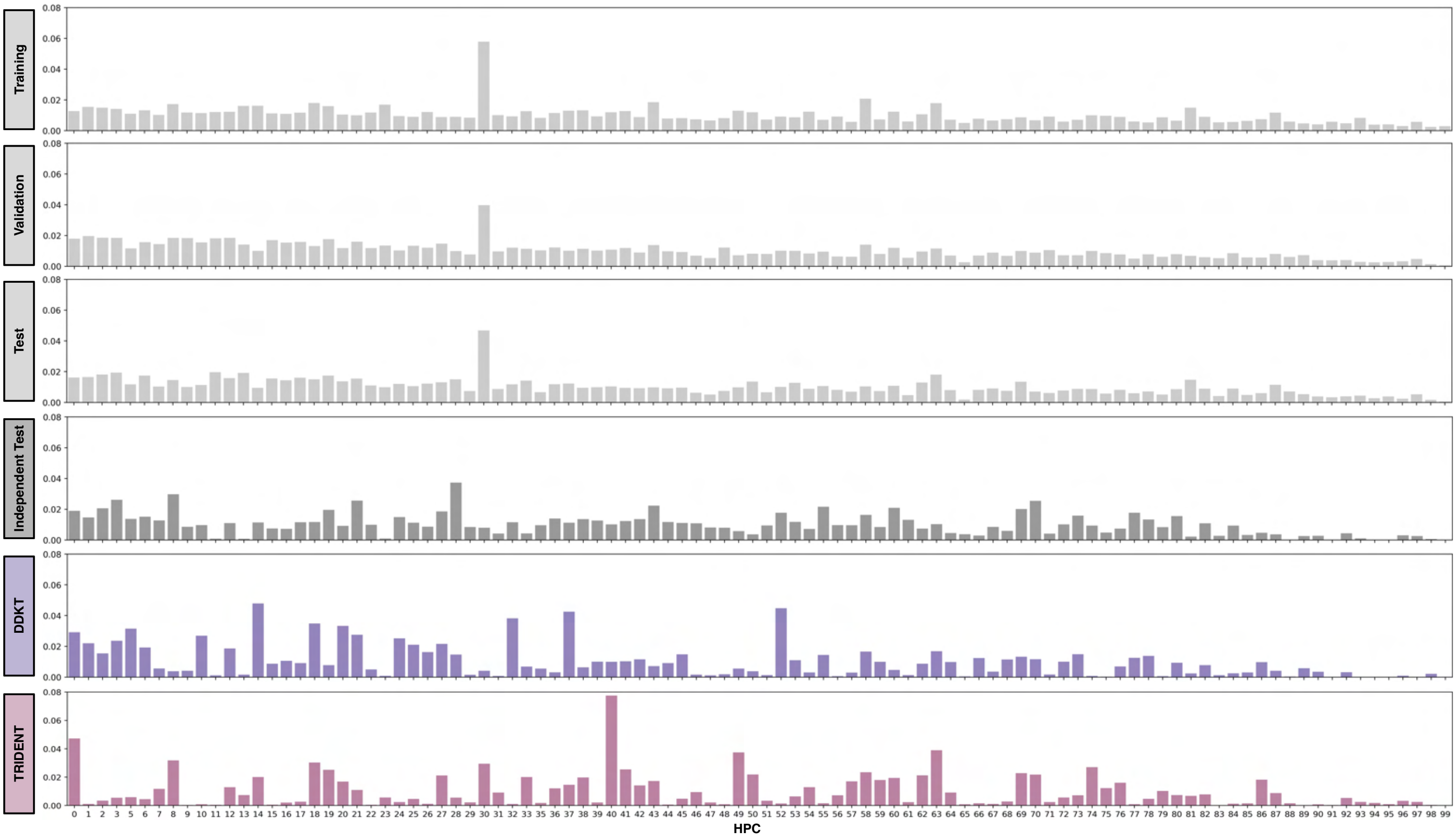


**Supplementary Fig. 3 | WSI-level distribution of HPCs at 10X magnification.** WSI vector representations were rescaled to exclude HPCs 4 and 34 (containing edge tiles). Average HPC presence (percent of WSI in a particular HPC) across patients plotted as a histogram by cohort.

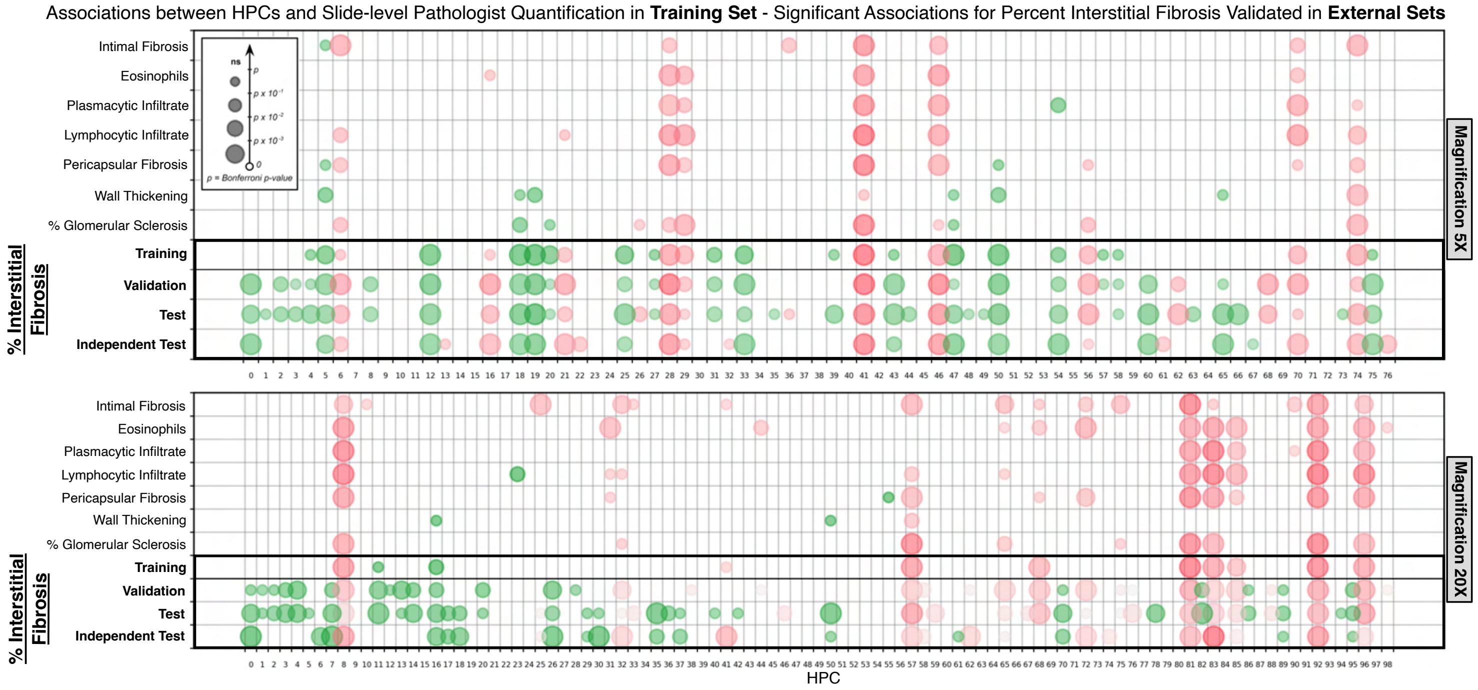


**Supplementary Fig. 4 | Analysis of HPC composition against expert pathologist quantification - 5X and 20X magnifications.** Associations between HPC composition and slide-level expert pathologist quantifications in the training set. Only associations passing Bonferroni threshold plotted as circles. Circle size increases with significance. Red circles indicate positive associations with slide-level pathology; green circles indicate negative associations with slide-level pathology. For HPCs significantly correlated with percent interstitial fibrosis in the training set, analysis was repeated in the validation, test, and independent test sets.

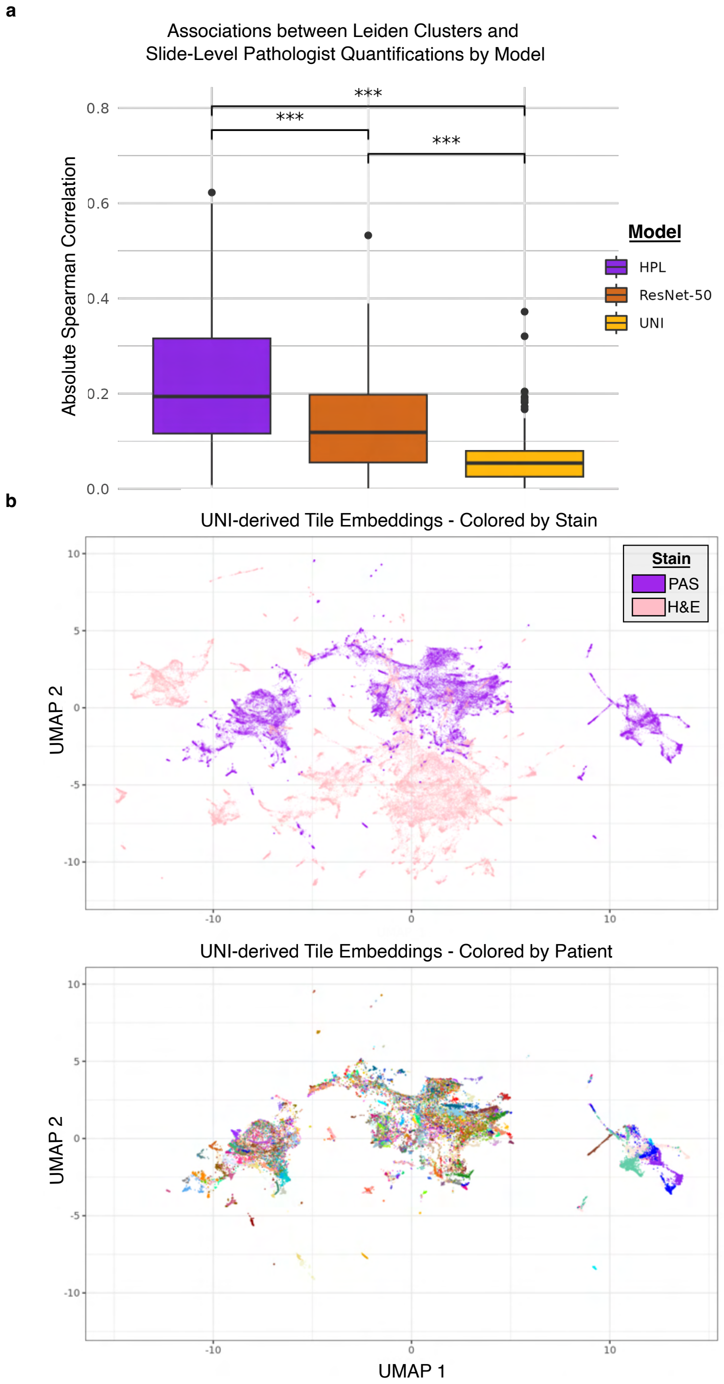


**Supplementary Fig. 5 | Comparative analysis of HPL against off-the-shelf SSL models. a,** Leiden clusters were defined using training, validation, and test set tile embeddings generated from pre-tained HPL, ResNet-50, and UNI. The absolute spearman correlations between the defined Leiden clusters and slide-level pathology are shown by model (*** indicates $p < 0.001$. **b,** In the TRIDENT set containing tissue stained with PAS and H&E, UNI-derived tile embeddings were visulized by stain and

**a**

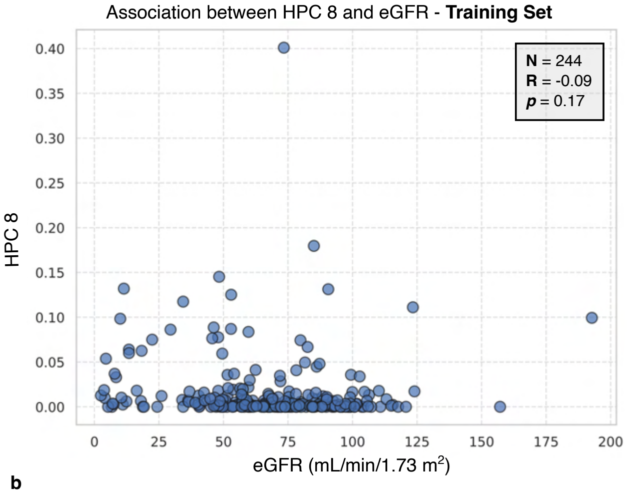


**b**

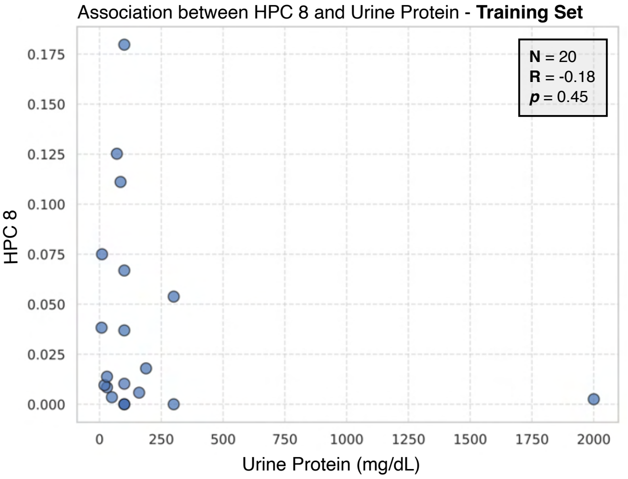


**Supplementary Fig. 6 | Investigating marker HPC 8 with medullar tissue patterns. a,** Associations between HPC 8 composition and estimated glomerular filtration rate in the training set. **b,** Associations between HPC 8 composition and urine protein in the training set.
