## Supplemental Table 1 for "Phenotypic and prognostic insights through unbiased self-supervised learning on kidney histology"

| | Overall | Patient Group 0 | Patient Group 1 | Patient Group 2 |
|---|---|---|---|---|
| **N** | 739 | 80 | 64 | 223 |
| **Age (yr)** | 60.8 (14.7) | 61.4 (16.8) | 59.9 (16.3) | 58.5 (14.0) |
| **Female** | 281 (38.0%) | 36 (45.0%) | 25 (39.1%) | 88 (39.5%) |
| **BMI** | 30.7 (8.2) | 30.3 (6.9) | 32.2 (8.7) | 31.6 (8.5) |
| **History of HTN** | 503 (68.5%) | 55 (69.6%) | 55 (85.9%) | 137 (61.7%) |
| **History of DM** | 261 (35.8%) | 23 (29.9%) | 22 (34.4%) | 77 (35.0%) |
| **Systolic Blood Pressure (mmHg)** | 136.0 (20.2) | 136.0 (20.0) | 137.7 (20.8) | 134.9 (18.6) |
| **Diastolic Blood Pressure (mmHg)** | 77.0 (12.2) | 78.7 (14.5) | 78.6 (11.3) | 76.9 (12.0) |
| **Serum Creatinine Level (mg/dL)** | 1.5 (1.7) | 1.4 (1.0) | 1.3 (1.3) | 1.1 (0.5) |
| **eGFR (CKD-EPI)** | 67.2 (27.4) | 62.1 (25.1) | 70.7 (25.6) | 75.6 (22.8) |
| **Blood Urea Nitrogen (mg/dL)** | 18.9 (10.1) | 20.8 (11.3) | 17.0 (7.7) | 16.6 (6.6) |
| **CKD stage** | 2.2 (1.2) | 2.4 (1.1) | 2.1 (1.1) | 1.9 (0.9) |
| **Intimal Fibrosis (0-3)** | 1.5 (0.9) | 1.1 (1.0) | 1.5 (0.8) | 1.2 (0.7) |
| **Eosinophils (0-3)** | 0.2 (0.5) | 0.2 (0.5) | 0.2 (0.4) | 0.1 (0.3) |
| **Plasmacytic Infiltrate (0-3)** | 0.4 (0.6) | 0.3 (0.5) | 0.3 (0.5) | 0.3 (0.5) |
| **Lymphocytic Infiltrate (0-3)** | 1.1 (0.8) | 0.9 (0.8) | 0.9 (0.8) | 0.9 (0.7) |
| **Pericapsular Fibrosis (0-2)** | 0.7 (0.7) | 0.7 (0.8) | 0.7 (0.6) | 0.5 (0.6) |
| **Wall Thickening (0-3)** | 0.2 (0.5) | 0.4 (0.7) | 0.1 (0.4) | 0.0 (0.2) |
| **Globally Sclerotic Glomeruli (%)** | 14.5 (20.6) | 15.1 (21.6) | 11.0 (11.3) | 8.3 (9.5) |
| **Interstitial Fibrosis (%)** | 13.4 (21.7) | 16.8 (24.6) | 7.2 (9.3) | 5.1 (5.2) |

**Supplementary Table 1. Analysis of patient groups defined using 10X HPC composition**

| Patient Group 3 | Patient Group 4 | p value | Not Missing |
|---|---|---|---|
| 128 | 244 | | 739 |
| 62.5 (12.5) | 62.0 (14.9) | **0.049** | 738 (99.9%) |
| 42 (32.8%) | 90 (36.9%) | 0.480 | 739 (100.0%) |
| 31.1 (8.2) | 29.4 (7.9) | **0.023** | 721 (97.6%) |
| 80 (63.0%) | 176 (72.7%) | **0.001** | 734 (99.3%) |
| 45 (35.2%) | 94 (39.2%) | 0.644 | 729 (98.6%) |
| 137.1 (21.8) | 136.2 (20.7) | 0.857 | 643 (87.0%) |
| 78.1 (12.6) | 75.6 (11.9) | 0.217 | 643 (87.0%) |
| 1.2 (0.6) | 2.2 (2.7) | **<0.001** | 736 (99.6%) |
| 70.2 (22.8) | 58.7 (31.7) | **<0.001** | 736 (99.6%) |
| 16.8 (7.6) | 22.1 (13.0) | **<0.001** | 710 (96.1%) |
| 2.1 (1.0) | 2.6 (1.3) | **<0.001** | 725 (98.1%) |
| 1.5 (0.8) | 1.9 (0.9) | **<0.001** | 682 (92.3%) |
| 0.3 (0.5) | 0.3 (0.5) | **<0.001** | 692 (93.6%) |
| 0.3 (0.6) | 0.7 (0.8) | **<0.001** | 692 (93.6%) |
| 0.9 (0.7) | 1.4 (1.0) | **<0.001** | 693 (93.8%) |
| 0.7 (0.7) | 0.9 (0.8) | **<0.001** | 692 (93.6%) |
| 0.2 (0.4) | 0.3 (0.7) | **<0.001** | 691 (93.5%) |
| 9.9 (9.7) | 23.9 (29.6) | **<0.001** | 695 (94.0%) |
| 8.2 (8.4) | 25.4 (31.3) | **<0.001** | 693 (93.8%) |
