## Supplemental Table 2 for "Phenotypic and prognostic insights through unbiased self-supervised learning on kidney histology"

| | Overall | Patient Group 0 | Patient Group 1 | Patient Group 2 |
|---|---|---|---|---|
| **N** | 738 | 72 | 124 | 176 |
| **Age (yr)** | 60.8 (14.6) | 61.4 (17.1) | 60.3 (17.4) | 60.1 (13.8) |
| **Female** | 281 (38.1%) | 32 (44.4%) | 49 (39.5%) | 65 (36.9%) |
| **BMI** | 30.6 (8.0) | 30.6 (7.1) | 27.8 (6.1) | 31.3 (8.1) |
| **History of HTN** | 503 (68.6%) | 48 (67.6%) | 98 (80.3%) | 122 (69.7%) |
| **History of DM** | 261 (35.9%) | 19 (27.5%) | 57 (47.5%) | 54 (30.7%) |
| **Systolic Blood Pressure (mmHg)** | 136.0 (20.2) | 135.3 (19.8) | 137.4 (22.3) | 135.1 (19.8) |
| **Diastolic Blood Pressure (mmHg)** | 77.0 (12.2) | 78.4 (15.1) | 76.0 (13.3) | 77.9 (11.5) |
| **Serum Creatinine Level (mg/dL)** | 1.5 (1.7) | 1.4 (0.9) | 2.9 (3.1) | 1.3 (1.4) |
| **eGFR (CKD-EPI)** | 67.1 (27.3) | 62.0 (25.0) | 51.5 (37.9) | 70.4 (23.6) |
| **Blood Urea Nitrogen (mg/dL)** | 18.9 (10.1) | 20.6 (11.6) | 24.9 (15.3) | 16.9 (7.7) |
| **CKD stage** | 2.2 (1.2) | 2.3 (1.1) | 3.0 (1.5) | 2.1 (1.0) |
| **Intimal Fibrosis (0-3)** | 1.5 (0.9) | 1.2 (1.0) | 2.0 (1.1) | 1.5 (0.8) |
| **Eosinophils (0-3)** | 0.2 (0.5) | 0.2 (0.5) | 0.4 (0.5) | 0.2 (0.5) |
| **Plasmacytic Infiltrate (0-3)** | 0.4 (0.6) | 0.3 (0.5) | 0.9 (0.9) | 0.3 (0.5) |
| **Lymphocytic Infiltrate (0-3)** | 1.1 (0.8) | 0.9 (0.8) | 1.8 (1.0) | 0.8 (0.8) |
| **Pericapsular Fibrosis (0-2)** | 0.7 (0.7) | 0.7 (0.7) | 1.2 (0.9) | 0.7 (0.6) |
| **Wall Thickening (0-3)** | 0.2 (0.5) | 0.3 (0.7) | 0.5 (0.9) | 0.2 (0.5) |
| **Globally Sclerotic Glomeruli (%)** | 14.5 (20.6) | 12.8 (17.7) | 38.2 (36.0) | 9.9 (11.8) |
| **Interstitial Fibrosis (%)** | 13.4 (21.7) | 14.6 (20.3) | 44.6 (35.9) | 7.6 (10.1) |

**Supplementary Table 2. Analysis of patient groups defined using 5X, 10X, and 20X HPC compos**

| Patient Group 3 | Patient Group 4 | p value | Not Missing |
|---|---|---|---|
| 185 | 181 | | 738 |
| 60.1 (13.3) | 62.5 (13.4) | 0.492 | 737 (99.9%) |
| 68 (36.8%) | 67 (37.0%) | 0.797 | 738 (100.0%) |
| 32.8 (9.1) | 29.7 (7.5) | **<0.001** | 720 (97.6%) |
| 129 (69.7%) | 106 (58.9%) | **0.003** | 733 (99.3%) |
| 69 (37.5%) | 62 (34.6%) | **0.021** | 728 (98.6%) |
| 135.9 (19.3) | 136.3 (20.3) | 0.922 | 643 (87.1%) |
| 77.2 (12.1) | 76.1 (11.4) | 0.551 | 643 (87.1%) |
| 1.1 (0.5) | 1.2 (1.3) | **<0.001** | 735 (99.6%) |
| 71.7 (21.5) | 72.0 (23.9) | **<0.001** | 735 (99.6%) |
| 17.7 (7.6) | 17.4 (7.9) | **<0.001** | 709 (96.1%) |
| 2.1 (0.9) | 2.0 (1.0) | **<0.001** | 724 (98.1%) |
| 1.4 (0.7) | 1.4 (0.8) | **<0.001** | 681 (92.3%) |
| 0.2 (0.4) | 0.2 (0.4) | **<0.001** | 691 (93.6%) |
| 0.4 (0.6) | 0.3 (0.6) | **<0.001** | 691 (93.6%) |
| 1.0 (0.7) | 0.9 (0.7) | **<0.001** | 692 (93.8%) |
| 0.6 (0.6) | 0.5 (0.6) | **<0.001** | 691 (93.6%) |
| 0.1 (0.3) | 0.0 (0.2) | **<0.001** | 690 (93.5%) |
| 10.8 (11.5) | 8.8 (10.2) | **<0.001** | 694 (94.0%) |
| 7.0 (7.7) | 6.7 (9.3) | **<0.001** | 692 (93.8%) |

**sition**
